# Brain charts harness population neuroscience to unlock neurodiversity of ADHD

**DOI:** 10.1101/2025.03.13.25323789

**Authors:** Ningning Liu, Yin-Shan Wang, Zi-Xuan Zhou, Peng Gao, Xinyi Zhang, Ziqing Zhu, Yuan Gao, Li-Zhen Chen, Haimei Li, Changxi Ju, Saashi Bedford, Clara Pecci Terroba, Richard A. I. Bethlehem, Yufeng Zang, Yufeng Wang, Lifespan Brain Chart Consortium, Chinese Color Nest Consortium, Lu Liu, Qiujin Qian, Xi-Nian Zuo

## Abstract

**Background:** Attention-deficit/hyperactivity disorder (ADHD) is a heterogeneous neurodevelopmental condition influenced by complex genetic and environmental factors, necessitating a population neuroscience approach to understand its etiology. Reclassifying ADHD subtypes through efficient and universally applicable measurements offers the potential to address its heterogeneity, thereby advancing personalized diagnosis and treatment.

**Methods:** This cross-sectional study leverages normative brain charts derived from 123,984 structural magnetic resonance images alongside extensive multimodal data – including blood, brain, behavioral, and environmental measurements – to unlock the neurodiversity of ADHD. We established centile scores of brain morphology in 270 children with ADHD (6-17 years) to identify deviations from the normative models adjusted from the lifespan brain charts. Using partitioning around medoids, ADHD individuals were clustered based on these deviations to identify trait constellations underlying ADHD’s clinical heterogeneity. Multidimensional data, including clinical symptoms, neurocognitive outcomes, brain function, and genetic and environmental risk factors, were compared to dissect the heterogeneity and support more customized early diagnosis or therapeutic approaches from a population neuroscience framework.

**Findings:** Regional deviations in cortical volume among children with ADHD were widely distributed, indicating pronounced individual differences. Further analysis revealed two subgroups of ADHD with distinct brain development patterns: delayed brain growth (DBG-ADHD) and precocious brain growth (PBG-ADHD). DBG-ADHD children exhibited significant neurocognitive impairments and higher functional homotopy within the default-mode network. Differentially expressed genes in this group were related to neurodevelopment, and more prenatal risk factors that affect brain development were identified. Conversely, PBG-ADHD was linked to elevated levels of disruptive behaviors, enhanced functional homotopy within language and somatomotor networks, and genetic pathways related to neurogenesis, while no significant prenatal environmental risk factors were identified.

**Interpretations:** Despite having similar core symptoms, ADHD encompasses two distinct subgroups characterized by fundamentally different developmental characteristics. One subgroup exhibited comprehensive developmental delays influenced by genetic and prenatal environmental factors. The other displayed congenital brain abnormalities that stem from atypical brain morphogenesis or surrounding unfriendly environmental factors, often associated with disruptive behaviors. These findings span a population neuroscience framework to provide actionable neurotargets to advance clinical practice through more tailored approaches.

**Funding:** The STI 2030 major projects (2021ZD0200500), the National Key R&D Program of China (2024YFC3308300), the National Natural Science Foundation of China (81220108014), the Capital’s Funds for Health Improvement and Research, the National Key Basic Research Program of China, the CAMS Innovation Fund for Medical Sciences, and the China Postdoctoral Science Foundation (2023M740147).

## Introduction

Attention-deficit/hyperactivity disorder (ADHD) is a common and highly heterogeneous neurodevelopmental disorder primarily influenced by genetic-environmental interactions.^1,2^ It affects approximately 5%^3^ to 7%^4^ of the global pediatric population. According to the Diagnostic and Statistical Manual of Mental Disorders, Fifth Edition (DSM-5), the symptoms of ADHD must be *inconsistent with the developmental level*.^5^ Clinical guidelines in various nations also underscore the importance of age-specific intervention strategies.^2^ However, current clinical practices are predominantly based on subjective assessments rather than objective, developmentally aligned criteria. This gap largely arises from a limited understanding of the neurodevelopmental mechanisms underlying ADHD.

Neuroimaging studies have sought to clarify these neurodevelopmental characteristics. Due to significant individual differences in ADHD, these studies face challenges in achieving clinical translation. Several longitudinal studies by Shaw et al. comparing ADHD children with typically developing controls (TDC) identified developmental delays in cortical thickness, surface area, gyrification, and cerebellar phenotypes.^6,7,8^ These differences are more pronounced during childhood and tend to diminish with age. However, subsequent small-scale studies have not consistently replicated these findings.^9,10,11,12,13^ Large-scale cross-sectional investigations, such as ENIGMA-ADHD,^14,15^ ADHD-200,^16^ and ABCD,^17^ have revealed similar developmental delays, although with effect sizes too small for clinical relevance. Recent studies have increasingly acknowledged that the potential heterogeneity of ADHD contributes significantly to the underlying problems.

Some studies have sought to classify the subtypes of ADHD according to underlying pathophysiological mechanisms, including symptoms, temperament, and cognition^18,19,20^ to advance clinical translation. However, these assessments are constrained by their behavioral focus and insufficient to uncover neurobiological underpinnings. In contrast, neuroimaging studies have employed magnetic resonance imaging (MRI) to assess tissue volume,^21^ cortical thickness,^22^ as well as functional activities of tasks^23^ and rest,^24,25,26^ or electroencephalogram (EEG),^27,28^ to classify ADHD into multiple subtypes characterized by different patterns of symptoms, behavior, emotion, cognition, gene expression profiles, or even treatment responses. These findings support the hypothesis that ADHD encompasses subtypes with distinct neurodevelopmental mechanisms, offering a promising avenue to address its inherent heterogeneity. The absence of reliable and globally consistent metrics for normative neurodevelopmental deviations has limited the generalization of such studies to the population level, hindering the identification of relevant mechanisms.

By integrating genetic and environmental data with population-level neuroimaging (Figure 1),^29,30^ this study aims to address these limitations. Our approach surpassed previous efforts (e.g.,^22,21,31^) in the robustness and generalizability of subtyping, and further revealed potential genetic and environmental pathways that contribute to individual variability in ADHD. To account for normative developmental changes and sex-specific variations in brain phenotypes, we introduced the emergent normative modeling approach to benchmark individual differences before subtyping. In contrast to a recent study,^22^ our integration of lifespan brain charts,^32^ derived from population-level neuroimaging data (*N* = 123, 984), enabled more robust and generalizable centile scores for normative neurodevelopmental deviations. This integration, leveraging population-level priors, significantly improved the external validity of our modeling outcomes.^33^ Beyond the relevance of symptoms, behavioral aspects and interhemispheric communication in imaging-derived ADHD subtypes (part of preliminary results have been reported in our previous presentation^34^), we integrated whole genome genotyping with environmental exposure data prenatal, perinatal and postpartum to elucidate the genetic and environmental origins of heterogeneity of ADHD. This comprehensive approach delineates an initial blueprint for understanding the neurodevelopmental mechanisms of ADHD.

**Figure 1.**
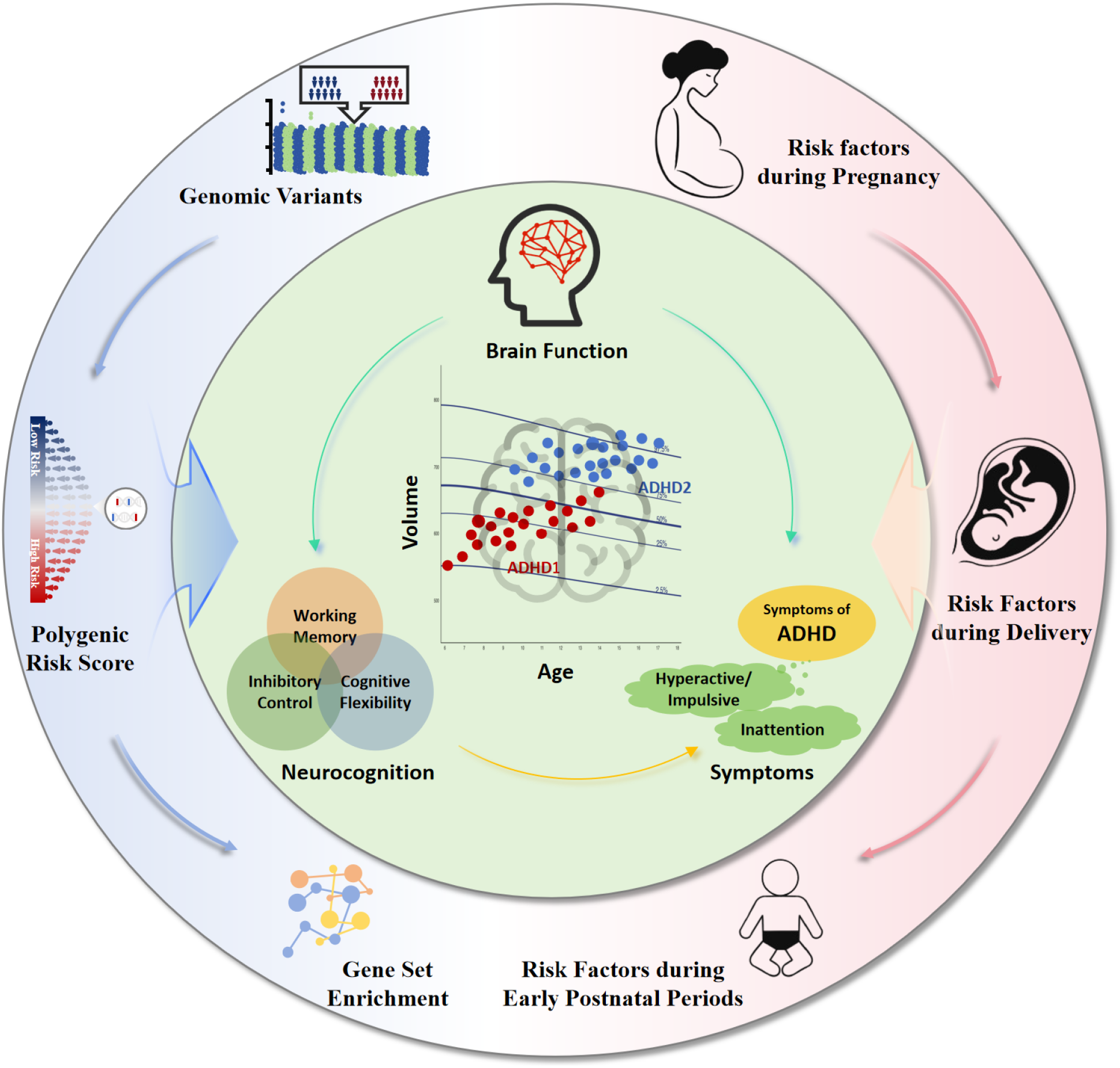
Population neuroscience research framework for unlocking the neurodiversity of ADHD with brain growth charts. Population neuroscience centers the human brain as a developmental outcome shaped by genetic and environmental processes and their interactions. Neuro-diversity is expressed through three facets: brain function, neurocognitive behavior, and clinical symptoms, derived from individuals with diverse growth patterns (indicated in different colors in the center) revealed by brain charts. As shown in the outward circle band, both genome-wide association (GWAS) and environment-wide association (EWAS) studies benchmark the heterogeneity of ADHD and parse its genetic (blue) and environmental (pink) links. GWAS identifies genomic variants, polygenic risk factors, and gene set enrichment, while EWAS delineates environmental risk factors during pregnancy and early postnatal periods.

### Research in context

**Evidence before this study**

We searched PubMed and Web of Science for studies on classifying ADHD subtypes using neuroimaging, covering all publications up to February 8, 2025. Our search strategy included the following terms in the title or abstract, without language restrictions: “Attention Deficit Disorder with Hyperactivity,” or “Minimal Brain Dysfunction,” and “subtypes,” “subgroups,” or “subtyping,” and “neuroimaging,” or “brain.” We identified 10 studies employing imaging modalities to perform data-driven clustering and identify biologically distinct subtypes of children with ADHD, independent of clinical diagnostic criteria. Although these studies support the existence of neurobiological subtypes, only one^22^ considered developmental effects on brain phenotypes and none investigated the genetic and environmental factors underlying differences in subtypes.

**Added value of this study**

We identified two new subgroups of ADHD with distinct neurodevelopmental characteristics: delayed brain growth (DBG-ADHD) and precocious brain growth (PBG-ADHD). Individuals diagnosed with DBG-ADHD exhibited reduced executive functioning and increased exposure to prenatal risk factors. Furthermore, this condition was associated with genetic pathways related to molecular signaling processes that are essential for developmental mechanisms. PBG-ADHD showed symptoms of disruptive behaviors and was associated with neurogenesis-related genetic pathways. These findings integrate genetic, environmental, and neurodevelopmental perspectives to elucidate the heterogeneity of ADHD.

**Implications of all the available evidence**

The typology derived from human brain charts offers a robust framework to address the heterogeneity of ADHD. Despite a uniform diagnosis, ADHD encompasses subgroups with different neurodevelopmental characteristics. One subgroup is characterized by multidimensional developmental delays, which can stem from altered brain development caused by innate genetic factors and prenatal environmental exposures. In contrast, another subgroup mainly exhibits precocious brain development and disruptive behaviors due to abnormal neurogenesis or unfriendly environmental factors. Validating these findings with other samples in future research holds promise for refining the nosology of ADHD and advancing personalized diagnostic and therapeutic strategies.

## Methods

### Participants and Clinical Measures

Data were collected from two sites in China using high-resolution 3.0 Tesla MRI scanners: Peking University Sixth Hospital/Institute of Mental Health (General Electric; Discovery MR750) and Beijing Normal University (Siemens; Trio Tim). Following rigorous data processing and filtering (Appendix page 1), the study included 270 patients with ADHD (6–16 years, 210 boys, 60 girls) and 127 typically developing children (54 boys, 73 girls) matched for cultural background. Participants were either stimulant-naive or had discontinued stimulant medication for at least 48 hours before evaluation to minimize medication effects.

All neuropsychological and neuropsychiatric tests were performed outside the MRI scanner. The caregivers most familiar with the children completed the assessments or interviews. All clinical and cognitive evaluations were performed at the Peking University Sixth Hospital/Institute of Mental Health by certified psychiatrists or psychiatric residents supervised by senior psychiatrists (Appendix pages 1–4). Written informed consent was obtained from the participants or their parents/guardians. The Research Ethics Review Board of Peking University Sixth Hospital approved the study procedures.

### MRI Data Processing

Each participant underwent T1-weighted volumetric (T1w) and functional MRI (fMRI) at rest. Detailed scan protocols, image pre-processing and quality control are provided in the Supplementary Material (Appendix pages 4– 6). All T1w and resting-state fMRI images were pre-processed using the Connectome Computation System.^35,36^ Six global volume phenotypes were analyzed: gray matter volume (GMV), white matter volume (WMV), subcortical gray matter volume (sGMV), ventricular cerebrospinal fluid volume (CSF), total surface area (totalSA), and global mean cortical thickness (meanCT). Regional analyses were performed on the volume of 34 bilateral gray matter regions, as defined by the Desikan–Killiany atlas.^37^ Bethlehem et al. noted that centile scoring based on their models is reliable for studies comprising scans *N >* 100. Due to T1-weighted data collected from two different MRI scanners, a harmonization technique named ComBat was performed on all 34 parcel volumes and six global MRI phenotypes to remove unwanted scan variability. The ComBat package in MATLAB was applied, accounting for sex and gestational age (birth date + 280 days) as biological variables, with local and global indices as variables of interest. Centile scores were generated for all individuals based on the human lifespan brain charts for all global and regional phenotypes of human brain morphology using Brent’s maximum likelihood estimation (see Appendix pages 5 for more details).

### DNA Extraction and Genotyping

Genomic DNA was isolated from peripheral blood using standard protocols (Omega Bio-tek Inc., Doraville, GA). All individuals had genotyping rates above 98%.

SNP genotyping and quality control are detailed in the supplementary material (see Appendix pages 7 for more details). The procedure of the genome-wide association studies (GWAS) used the package “rMVP” in R (v4.0.3), applying rigorous quality control using PLINK 1.9. Genome-wide association analyses were performed using PLINK software with three genetic models to identify SNPs associated with phenotypes. The fixed and random model Circulating Probability Unification (FarmCPU) was used for parallel-accelerated association tests due to its higher statistical power than other methods (e.g., GLM and MLM).^38^ The ADHD polygenic risk score (PRS) was derived from the GWAS findings in our previous research, using a discovery cohort that included 1033 cases of ADHD and 950 healthy controls (total N=1,983). This is the largest Han Chinese population sample investigated for genetic risk of ADHD to date. Details of the collection dataset can be found in a previous study.^39^

### Environmental Risk Factor Assessment

Environmental risk factors were assessed using the Handbook on the Living Environment of Children and Adolescents. Given the critical periods identified for environmental events associated with ADHD,^2^ our main analysis focused on pregnancy, delivery, and early postnatal periods (see Appendix pages 8 for full details).

### Statistical Analysis

#### Deviation of brain development relative to the normative charts

Baseline characteristics of ADHD and TDC were summarized using descriptive statistics for continuous and categorical measures (Table 1). Centile scores were generated for all individuals based on the human lifespan brain charts for all global and regional phenotypes of human brain morphology using Brent’s maximum likelihood estimation. First, we evaluated group-level differences in the raw volume following conventional case-control analysis. Next, we compared logarithmically transformed centile scores between ADHD and TDC using a two-sample t-test. Subsequently, to identify biology–psychopathology associations, we assessed correlations between ADHD behaviors and raw data or individual deviations (see Appendix pages 8–9 for more details).

**Table 1:** Demographic and Clinical Characteristics of the Participants. The subtype of ADHD was performed using the Clinical Diagnostic Interview Scale (CDIS); IA = Inattentive subtype; HI= Hyperactive and impulsive subtype; C = Combined subtype. The comorbidity of ADHD was performed using the Clinical Diagnostic Interview Scale (CDIS); ODD = oppositional defiant disorder; CD = conduct disorders; Anxiety disorders = specific phobia, social phobia, separation anxiety disorder, generalized anxiety disorder and obsessive compulsive; Mood disorders = dysthymic disorder, major depressive disorder, and bipolar affective disorder; Total IQ score was assessed by Chinese Wechsler Intelligence Scale III (WISC-III), with higher scores indicating greater cognitive abilities. One child’s full-scale IQ was assessed with the Wechsler Adult Intelligence Scale-Third Edition as he was 17 years old when he participated in the assessment. On the ADHD Rating Scale-IV (ADHD RS-IV), the range of the inattention scores is from 0 to 27, with higher scores indicating more attention. The range of the hyperactivity/impulsivity scores is from 0 to 27, with higher scores indicating more hyperactivity/impulsivity problems. The total ADHD score was calculated as the sum of the inattention and hyperactivity/impulsivity scores. Scores range from 0 to 54. A greater score indexes more severe ADHD symptoms. Student’s t-test was used for the statistical analyses, except for the total score of ADHD RS-IV, for which the Mann-Whitney U test was used.

| Characteristic | ADHD (n=270) | TDC (n=127) | p-Value |
| --- | --- | --- | --- |
| Month - m | 124.08±29.99 | 129.04±23.11 | 0.070 |
| Sex - no. (%) |  |  | < 0.001 |
| Boy | 210 (77.8) | 54 (42.5) | .. |
| Girl | 60 (22.2) | 73 (57.5) | .. |
| Stimulant medication - no. (%) | 7 (2.6) | / | .. |
| ADHD subtype - no. (%) |  |  | .. |
| IA | 170 (63.0) | / | .. |
| HI | 2 (0.7) | / | .. |
| C | 98 (36.3) | / | .. |
| Comorbidity - no. (%) |  |  | .. |
| ODD | 49 (18.1) | / | .. |
| CD | 3 (1.1) | / | .. |
| Tic disorder | 9 (3.3) | / | .. |
| Anxiety disorder | 1 (0.4) | / | .. |
| Mood disorder | 3 (1.1) | / | .. |
| Total IQ score | 103.72±14.08 | 118.57±12.35 | < 0.001 |
| ADHD RS-IV |  |  |  |
| Inattentive | 18.12±3.71 | 8.60±4.28 | < 0.001 |
| Hyperactive/impulsive | 11.55±6.20 | 6.00±4.20 | < 0.001 |
| Total | 29.67±7.94 | 14.73±7.45 | < 0.001 |

#### Defining new ADHD subgroups with different brain development patterns in brain charts

To examine whether individual deviations could better parse the heterogeneity of ADHD, we used partitioning around medoids (PAM), a robust clustering technique that uses a dissimilarity matrix for partitioning,^40^ to identify groups based on centile scores. The overall silhouette coefficient, which represents the average value across all points, serves as the criterion for selecting the most coherent and distinct clustering solution as the final result (Appendix pages 8–9).

#### Key Clinical Characteristics Difference of ADHD Subgroups

We conducted pairwise comparative analyses to determine if the identified ADHD subtypes represented clinically distinct subgroups compared to the TDC. Each ADHD subgroup was individually matched with the TDC participants by sex and age through manual matching to ensure comparability between groups, considering that cognition varies continuously with age during childhood and adolescence.

For each ADHD subgroup-TDC pair, we employed independent t-tests for continuous variables in the following analyses. The variables examined included the severity of the core symptoms of ADHD (inattention, hyperactivity-impulsivity, and total scores), comorbidity measures (oppositional defiant disorder, conduct disorder, and anxiety/depression scores), and components of executive function (working memory, cognitive flexibility, and inhibitory control). Statistical significance was established at *p <* 0.05. Statistical analyses were performed with R v4.1.3 (IBM Corp., Armonk, NY, USA).

#### Homotopic functional affinity calculation

Due to the unequal sample sizes observed between the DBG-ADHD/PBG-ADHD and TDC groups (Appendix pages 13), to achieve demographically matched samples between the two ADHD subgroups and the TDC group, we used data from the development stage of the Chinese Color Nest Project (devCCNP), which is being developed as a valuable resource^41,42^ and has contributed to brain charts for the human lifespan.^35,32^ By combining the data from the TDC group with the devCCNP dataset, age and sex matching procedures were performed separately for the DBG-ADHD and PBG-ADHD groups. Functional Homotopic Affinity (HFA) represents a robust metric to assess the reliability and validity of functional integration and regional specificity within homotopic brain areas. In contrast to conventional methodologies, HFA facilitates a more precise quantification of inter-individual variability. This metric provides critical information on the intricate interplay between functional integration and regional specialization across cerebral structures, with particular emphasis on higher-order cognitive domains^43^(Appendix page 6). Consequently, HFA calculations were performed independently for both the DBG-ADHD versus TDC1 cohorts and the PBG-ADHD versus TDC2 groups. A higher functional homotopic affinity reflects greater functional homogeneity between homotopic vertices, while a lower affinity indicates reduced homogeneity and increased functional heterogeneity between these vertices.^44^ Furthermore, based on segmentation of the functional brain network parcellation, DU15NET,^45^ the current study calculated the effect sizes for various subregions within the network.

#### GWAS for identifying genetic variants and biological processes in subgroups of ADHD

A one-way analysis of variance (ANOVA) was performed to assess the differences in PRS between subgroups of ADHD and TDC. The associations between PRS and the centile score for all volumes were also calculated separately for the two subgroups of ADHD. The protein-protein interaction (PPI) network analysis was performed using the STRING database (v12.0; http://string-db.org). We used the multiple protein input option, maintaining all default settings except for restricting interactions to those with the highest confidence scores (* 0.900). To improve clarity, disconnected nodes were excluded from the network output. To further explore whether particular Gene Ontology (GO) terms occur more frequently than expected by chance in different subtypes, GO biological processes pathway enrichment analysis was conducted using STRING. This prioritized Biological Processes (BP) in GO to elucidate the molecular mechanisms underlying the phenotypic differences. The top five enriched GO terms were plotted.

#### Environment-Wide Associations of ADHD Subgroups

Due to the insufficient data on environmental exposure risk factors for TDC, we directly compared the differential contribution of environmental risk factors between ADHD subgroups. Specifically, between-group comparisons were conducted using independent samples t tests for continuous variables and Pearson’s Chi-square tests for categorical variables to assess differences in environmental risk factors between the two ADHD subgroups.

#### Sensitivity analysis

To make meaningful comparisons with the existing literature, we perform both cluster analysis and validation of clinical indicators with an identical analytical framework but with raw data. To verify the robustness of our findings, we used original individual centile scores, unadjusted by logarithmic transformation, to replicate cluster analysis.

## Results

### Participant Characteristics and Group Differences between ADHD and TDC Individuals

The demographic and clinical characteristics of the participants are detailed in Table 1. After applying site-effect harmonization using ComBat, neuroimaging analysis revealed no significant differences between the ADHD and TDC groups in six global metrics or 34 GMV parcels when using raw brain volume data, following Bonferroni corrections (|Cohen’s *d*| : 0.004 – 0.260; Figure S1A). However, a distinct pattern emerged when analyzing centile scores. The ADHD group showed significantly lower centile scores in four global metrics (WMV, sGMV, GMV, and totalSA) and 18 of 34 GMV parcels after Bonferroni corrections (|Cohen’s *d*| : 0.084 – 0.524; Figure S1B). The most affected regions were located mainly in the frontal and temporal lobes, including the superior frontal (|Cohen’s *d*| = 0.435), inferior temporal (|Cohen’s *d*| = 0.410), and supramarginal (|Cohen’s *d*| = 0.407) (see Appendix pages 10–11 for more details). These findings suggest a stunted developmental trajectory in specific brain regions for individuals with ADHD (Figure S1).

Although raw brain volume data did not show significant associations with behavioral measures, centile scores revealed a notable pattern: Individual centile scores of brain morphology were weakly correlated with the severity of ADHD symptoms, but were significantly associated with executive functions (EF), such as digit span backward (DSB), shift time, color interference time (IC), and word interference time (IW). Higher centile scores were significantly correlated with improved executive function performance in the ADHD cohort (Figure S1).

### Redefine ADHD into Subgroups Reflecting Distinct Brain Developmental Patterns

Individual centile scores of brain structures demonstrated broad variability in both the ADHD and TDC groups. However, the ADHD group showed a marked trend towards lower centile values, indicating a higher prevalence of infra-normal measurements (Figure S2). Cluster analysis revealed two subgroups of ADHD with distinct brain developmental patterns (Figure 2A): delayed brain growth (DBG-ADHD) and precocious brain growth (PBG-ADHD) (Demographic and clinical characteristics see Appendix pages 14).^46^ The brain growth centile of the DBG-ADHD group was significantly lower, and most individuals did not exceed the 50th percentile of the general population of the same age and sex. In contrast, individuals in the PBG-ADHD group exhibited significantly higher brain growth centiles compared to the general population.

**Figure 2.**
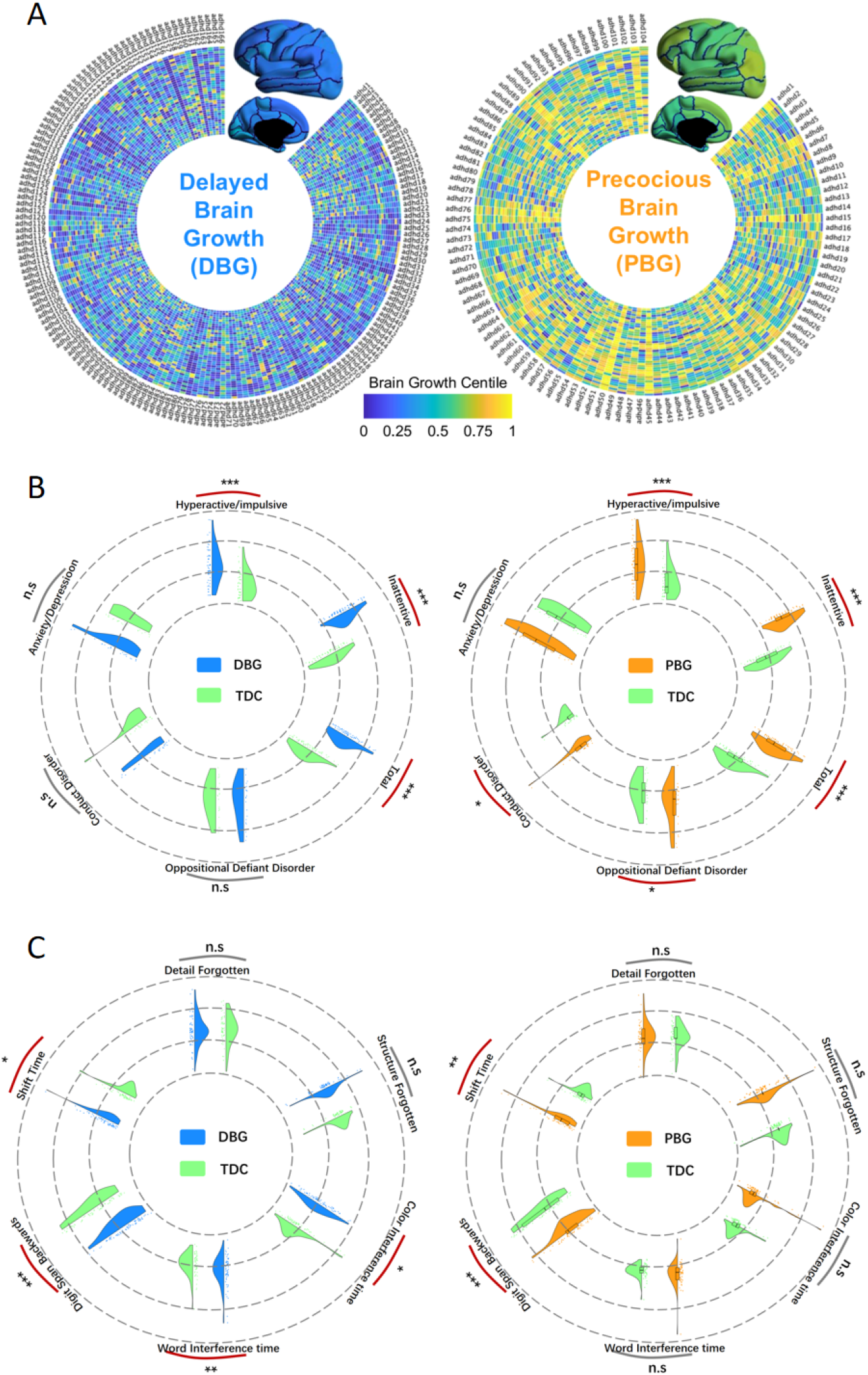
Brain developmental patterns and clinical characteristics in DBG-ADHD and PBG-ADHD subtypes. **[A]** Brain growth centile distribution maps showing regional maturation patterns in DBG-ADHD and PBG-ADHD groups (brighter colors indicate more mature regions). Differential symptom profiles between DBG-ADHD vs. TDC (left panel) and PBG-ADHD vs. TDC (right panel). **[C]** Executive function differences comparing DBG-ADHD vs. TDC (left panel) and PBG-ADHD vs. TDC (right panel). Red lines indicate statistical significance (^*^*p <* 0.05; ^**^*p <* 0.01; ^***^*p <* 0.001). DBG = Delayed brain growth group; PBG = Precocious brain growth group; TDC = Typically developing control.

**Figure 3.**
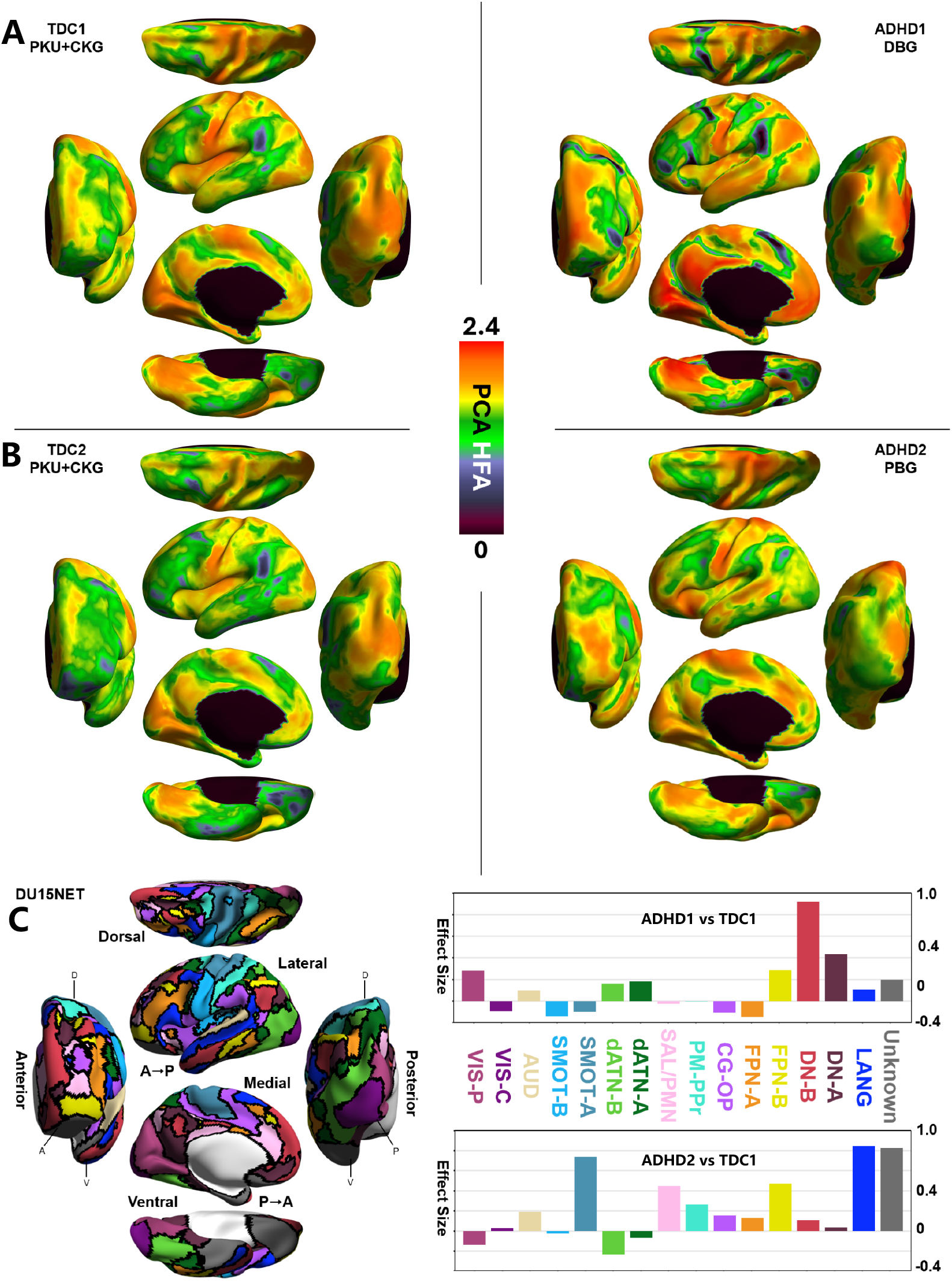
Homotopic functional affinity of two ADHD subgroups and healthy control. The warmer colors represent higher HFA values, while cooler colors indicate lower HFA values. **[A]** presents the whole-brain HFA map for the DBG-ADHD vs. TDC; **[B]** presents the whole-brain HFA map for the PBG-ADHD vs. TDC; **[C]** presents the effect size values for various subregions of the DU15NET functional brain network, highlighting the comparisons between the two ADHD subgroups and the TDC group. VIS-P: Visual Peripheral; VIS-C: Visual Central; AUD: Auditory; SMOT-B: Somatomotor-B; SMOT-A: Somatomotor-A; dATN-B: Dorsal Attention-B; dATN-A: Dorsal Attention-A; SAL/PMN: Salience/Parietal Memory Network; PM-PPr: Premotor-Posterior Parietal Rostral; CG-OP: Cingulo-Opercular; FPN-A: Frontoparietal Network-A; FPN-B: Frontoparietal Network-B; DN-B: Default Network-B; DN-A: Default Network-A; LANG: Language.

### Different Clinical Features Across the Two Subgroups

Compared to the TDC group, both DBG-ADHD and PBG-ADHD exhibited higher scores for the core symptoms of ADHD (*p <* 0.001). However, no significant differences were observed in the core ADHD symptoms scores between the two groups (*p* = 0.890). Furthermore, PBG-ADHD showed a higher ODD (*p <* 0.05) and CD score (*p <* 0.05) compared to TDC, while no differences were found between the DBG-ADHD and TDC groups.

Upon evaluating EF in three distinct groups, it was observed that there were no significant differences in performance on the visual working memory test (RCFT) (*p >* 0.05). However, individuals in DBG-ADHD exhibited significantly inferior performance compared to the TDC group in auditory working memory (DSB,*p <* 0.001), inhibitory control (Stroop test, *p <* 0.05), and cognitive flexibility (TMT) (*p <* 0.05). Children in the PBG-ADHD group exhibited worse auditory working memory performance (*p <* 0.001) and cognitive flexibility (*p <* 0.01) but not inhibitory control (*p >* 0.05).

To validate the results above, the above analysis is repeated with the data unmatched. The results were essentially unchanged appreciably (Appendix pages 14–15).

### Interhemispheric Communication across the ADHD Subgroups and TDC

Demographic information is available on Appendix pages 15–16. DBD-ADHD group mainly exhibits higher HFA values than the TDC1 group, particularly in the occipital and temporal lobes. Specifically, significant differences in HFA values were observed between the DBD-ADHD and TDC groups within the default network, with the most pronounced disparity occurring in the subregion B, where the effect size approaches a value of 1. In addition, the effect size for the difference between the two groups exceeds 0.4 in the Default Network A subregion.

The PBD group shows elevated HFA values, particularly in the prefrontal cortex, temporal lobe, occipital lobe, and regions adjacent to the central sulcus. When examining functional network subregions, the most substantial effect size is observed in the Language network(greater than 0.8), followed by Somatomotor-A (greater than 0.6).

### Neurobiological Profiles Across the Two Subgroups

Utilizing a significance threshold of 10^−6^, the GWAS identified five loci (ERBB4, FERD3L, TWISTNB, PCDH10, PABPC4L) associated with the linkage between DBG-ADHD and TDC. In addition, a singular locus, AUTS2, was found in the correlation between PBG-ADHD and TDC. Our analysis did not uncover any shared risk variants between DBG-ADHD and PBG-ADHD (Fig. 4A).

**Figure 4.**
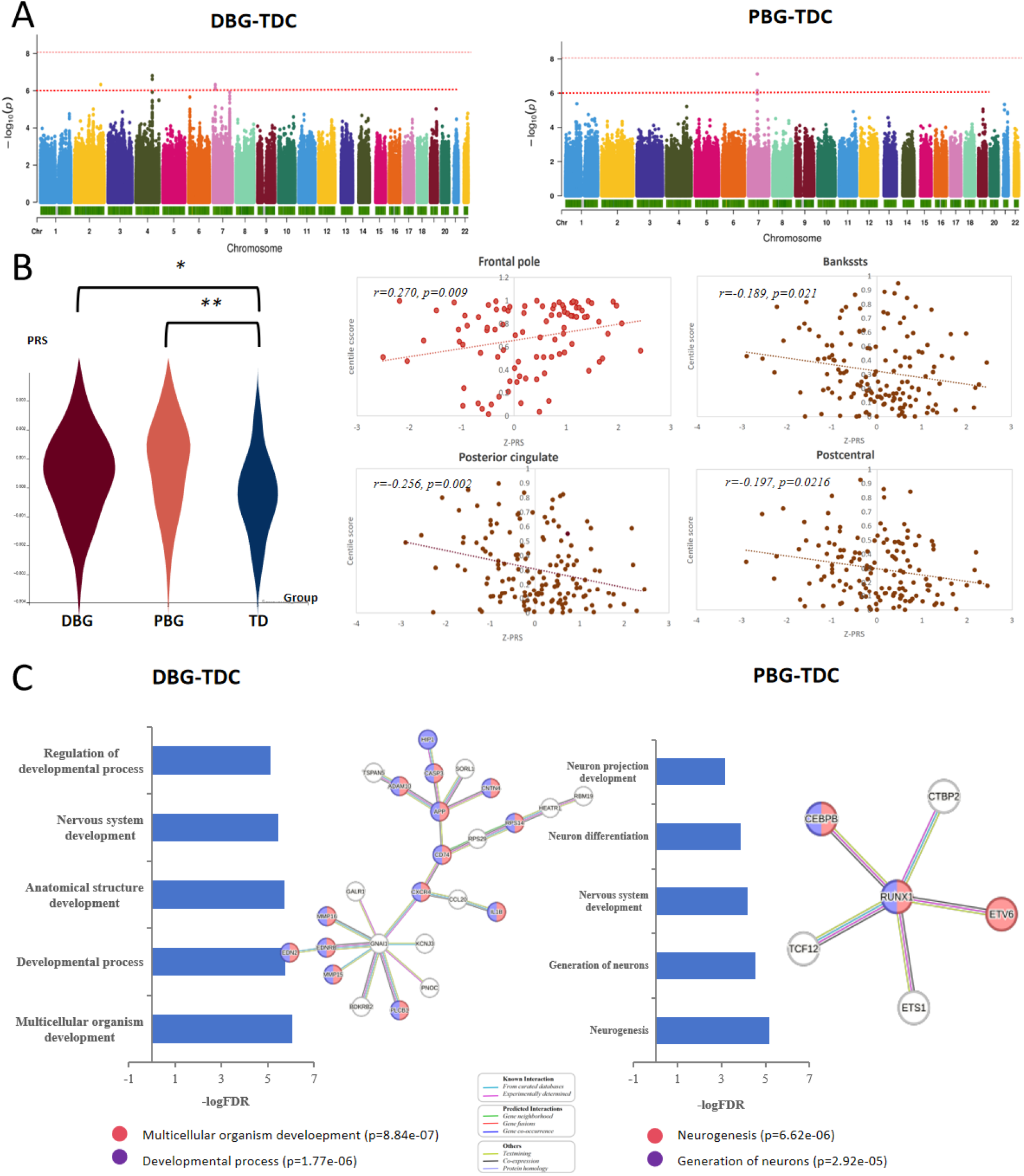
Distinct neurobiological profiles of DBG-ADHD and PBG-ADHD. **[A]** Results of the Genome-Wide Association Study (GWAS) for DBG-ADHD and PBG-ADHD. **[B]** Left: Comparison of Polygenic Risk Scores (PRS) among DBG-ADHD, PBG-ADHD, and TDC groups. Right: Scatter plots illustrating the relationship between PRS and centile scores of structural MRI metrics for DBG-ADHD and PBG-ADHD groups (Orange: DBG-ADHD; Brown: PBG-ADHD). **[C]** Gene Ontology (GO) enrichment analysis and Protein-Protein Interaction (PPI) network analysis for DBG-ADHD vs. TDC and PBG-ADHD vs. TDC comparisons. DBG = delayed brain growth group; PBG = precocious brain growth group; TDC = typically developing control.

Both DBG-ADHD (*p* = 0.012) and PBG-ADHD (p¡0.001) were characterized by a higher PRS score, although there were no significant differences between DBG-ADHD and PBG-ADHD (p=0.124). PRS was negatively correlated with centile scores for bankssts (r = -0.189; p = 0.021), postcentral (*r* = − 0.197; *p* = 0.016), and posterior cingulate (r = -0.256; p = 0.002) in the DBG-ADHD group, indicating that a higher genetic risk was associated with more severe delays in brain development. In contrast, PRS was positively correlated with centile scores for the frontal pole (r =0.270; p = 0.009) in the PBG-ADHD group, suggesting an association with precocious brain development. These findings align with their characteristic brain development patterns (Fig. 4B).

PPI analysis revealed a significant enrichment of known interactions between DBG-ADHD and TDC (*p* = 0.00136) and between PBG-ADHD and TDC (*p* = 0.0099). In the GO analysis, the BP analysis was performed to elucidate the functional implications of differentially expressed genes (DEGs) between the DBG-ADHD and the TDC group. The results of the BP analysis revealed a significant enrichment of these genes in several key terms related to the broader developmental and morphogenetic processes. These prominently represented terms include “Multicellular organism development” (false discovery rate [FDR]= 8.84 × 10^−7^), “Developmental process” (FDR= 1.77 × 10^−6^), “Anatomical structure development” (FDR= 1.90 × 10^−6^), “Nervous system development” (FDR= 3.48 × 10^−6^), and “Regulation of developmental process” (FDR= 7.85 × 10^−6^). Subsequently, the DEGs between the PBG-ADHD and TDC groups were analyzed. In contrast to the DBG-ADHD group, the PBG-ADHD group showed relatively lower significance levels. BPs predominantly related to “neurogenesis” (FDR= 6.62 × 10^−6^), “Generation of neurons” (FDR= 2.92 × 10^−5^), “Nervous system development” (FDR= 6.21 × 10^−5^), “Neuron differentiation” (FDR= 1.40 × 10^−4^), and “Neuron projection development” (FDR= 7.00 × 10^−4^), highlighting a more neurogenesis-specific enrichment pattern (Fig 4C).

### Environment-wide association of the ADHD Subgroups

No differences were apparent between the two groups of ADHD in terms of environmental risk factors during delivery or early postnatal periods (*p >* 0.05). However, significant differences during pregnancy included lower birth weights (3174.93 *±*689.13 g vs. 3603.29*±* 902.58 g), higher rates of preconceptual paternal alcohol habit (31. 9% versus 16. 7%), and greater exposure to maternal and passive smoking during pregnancy (36. 6% versus 15. 7%) in the DBG-ADHD group compared to PBG-ADHD.

#### Sensitivity analysis

The characteristics of the ADHD subgroups defined by the raw data were also provided in the supplemental material, to better understand the results of this study. After analyzing the raw data, we found that the differences between the two groups were minimal in clinical characteristics, cognitive characteristics, GWAS, and EWAS analyzes (Appendix pages 16–17). With the original individual centile scores, we did not find significant changes in the results.

## Discussion

To our knowledge, this was the first dissection of the developmental heterogeneity of ADHD with human lifespan brain charts and further revealed the potential genetic and environmental factors that contribute to individual variability in ADHD within a population neuroscience framework. Despite the same core symptoms, we found that children with ADHD in the current psychiatric diagnostic system were driven by two different developmental patterns that stemmed from different genetic and environmental factors, which corresponded to different clinical presentations, neurocognitive profiles, and brain functions.

The observed heterogeneity within the ADHD population in previous studies highlights the need for a paradigm shift toward identifying subgroups with divergent etiologies and addressing limitations in traditional psychiatric nosology. However, progress is hindered by the lack of methodologies that explicitly accommodate potential clinical sample heterogeneity. In particular, the objective of redefining the ADHD subgroups is to establish a unified approach globally that improves the practicality of diagnosing and treating ADHD. Thus, the methodologies used for subgroup classification should be straightforward and generalizable to all datasets, to facilitate future clinical applications. However, due to the absence of a consistent method worldwide, previous research on the heterogeneity of ADHD often remains specific to the datasets used, making it challenging to compare findings between studies and generalize results. Using the normative brain charts, our study performed cluster analyses to delineate two new subsets that have the potential to become recognized clinical diagnostic categories. Our analysis identified two distinct subgroups: DBG-ADHD, marked by delayed brain growth, consistent with findings from previous studies on ADHD, and PBG-ADHD, characterized by precocious brain growth. The contrasting characteristics of the two subgroups would explain the prior inconsistencies in the literature, as it appears to be the DBG-ADHD subgroup that drives the intergroup differences reported in previous case-control studies, which observed developmental delays in a wide variety of brain regions in ADHD.^6, 14, 15, 17^

Interestingly, core ADHD symptoms were consistent across both subgroups, underscoring the limitations of current symptom-based diagnostic checklists in distinguishing between these etiologically distinct groups. However, compared to TDC, the DBG-ADHD subgroup exhibited comprehensive deficits in executive function, while the PBG-ADHD subgroup displayed more pronounced symptoms of disruptive behavior disorders. This finding is consistent with our previous research, which indicated a positive correlation between more severe ODD/CD symptoms and executive function in ADHD, particularly in tasks that require inhibitory control, such as the Stroop task.^13^ Importantly, children with comorbid ODD/CD tend to show more mature brain function patterns resembling adult ADHD.^47^ Based on these robust findings, we hypothesize that DBG-ADHD represents a correctable developmental delay. Conversely, PBG-ADHD seems to constitute a developmental deviation characterized by compensatory mechanisms, which may pose greater challenges for intervention and whose symptoms are more likely to persist into adolescence.

In terms of brain function, our findings revealed higher HFA values in the DMN for the DBG-ADHD group, consistent with its role as an ‘intrinsic’ system often implicated in the pathophysiology of ADHD. This further suggests that DBG-ADHD represents a group that was previously described in the literature as exhibiting abnormalities in DMN.^48,49,50^ In addition, the damaged areas of the brain correspond to comprehensive deficits in executive function observed in this group. In contrast, the PBG-ADHD group shows primarily abnormalities in the SMOT and LANG brain networks, which are responsible for simpler tasks and mature earlier than the DMN. The network abnormalities observed in PBG-ADHD may, to some extent, explain their ODD/CD behaviors.

Then what shapes the difference between DBG-ADHD and PBG-ADHD? We explored this question from a population neuroscience framework, considering both the GWAS and EWAS analyses. Compared to the PBG-ADHD and TDC groups, DBG-ADHD exhibited more pronounced genetic differences, with enriched pathways linked to developmental processes, suggesting a strong genetic contribution to delayed brain growth. In contrast, the PBG-ADHD genes showed lower enrichment and were primarily related to neurogenesis rather than to broader developmental processes. Previous studies of common and rare variants have highlighted risk genes related to brain development in ADHD.^2,51^ Our results again suggest that this conclusion applies specifically to the DBG-ADHD subgroup. Regarding environmental risk factors, DBG-ADHD was relatively more exposed to prenatal risk factors, including low birth weight, preconceptual paternal alcohol habits, and passive maternal tobacco exposure during pregnancy. These prenatal factors can affect gene expression and brain development in offspring, increasing susceptibility to ADHD..^52,53,54^

These results demonstrate that the DBG-ADHD subgroup appears to originate from innate brain developmental abnormalities. Interestingly, individuals in the PBG-ADHD subgroup exhibited advanced brain morphology, poorer functional differentiation of the brain in certain networks responsible for simpler tasks, and more symptoms of ODD/CD. Importantly, the differentially expressed genes in this group demonstrate neurogenesis-related specificity, rather than strong neurodevelopmental specificity. This suggests that deviations from typical brain development may play a significant role in this process. Of course, we do not track and explore the environmental factors or current health conditions of the two groups. Recent longitudinal studies on ADHD suggest that symptoms of ADHD fluctuate, possibly in response to environmental or health-related factors.^55^ Therefore, PBG-ADHD, which has lower levels of differentially expressed genes and the absence of significant perinatal risk factors, may also be due to current environmental influences or health conditions. Regardless of which hypothesis is correct, DBG-ADHD can respond more effectively to pharmacological interventions targeting biological factors, while PBG-ADHD could benefit from non-pharmacological therapies addressing behavioral and social impairments. It should be noted that previous research has found that medications, especially stimulants of the central nervous system, can improve the core symptoms of ADHD, and the underlying mechanism may be the correction of the delayed brain development associated with ADHD by drugs.^56,57^ Similarly, in the genetic realm, studies have found that ADHD treatment medications affect genes and molecular processes related to neurodevelopment, thus promoting the normalization of brain function.^58,59^ This suggests that compared to PBG-ADHD, DBG-ADHD could respond better to medication.^60^ Future studies should examine these hypotheses in more detail to guide personalized ADHD treatment strategies.

This study has some limitations. First, it focused on an Asian population, and future studies are needed in diverse populations to validate these findings and advance global standardization of the diagnosis and treatment of ADHD. Second, as previously mentioned, we cannot explore whether the centile score or clusters of ADHD are associated with different prognoses or treatment responses, since we have not yet completed the follow-up of these children with ADHD. Future longitudinal studies should investigate treatment responses and outcomes in ADHD subgroups, particularly exploring hypothesized differential responsiveness to pharmacological and behavioral interventions.

In conclusion, the cluster analysis using individual centile scores identified two distinct subgroups of ADHD. DBG-ADHD exhibited delayed brain growth and poorer EF, possibly due to innate developmental delays. In contrast, PBG-ADHD demonstrated precocious brain growth and behavioral problems, likely influenced by deviations in normal brain genesis or environmental factors. This study implies that ADHD, as currently diagnosed based on clinical symptoms, represents fundamentally different subgroups. Our results provide valuable information for the future improvement of the ADHD diagnostic system and the individualized treatment of ADHD.

## Data Availability

All data produced in the present study are available upon reasonable request to the authors.

https://ccnp.scidb.cn/en

http://www.brainchart.io

## Contributors

NNL, YSW, ZXZ, and PG drafted the manuscript. All authors revised the manuscript. NNL and YSW performed data pre-processing and analysis. PG analyzed the fMRI data. ZQZ performed the genetic PRS data analysis. XYZ was responsible for searching for the literature review. HML managed the data. YG contributed to the preparation of the figures. LZC participated in the pre-processing of functional data. XNZ, QJQ, and LL served as project managers and reviewed the manuscript. XNZ, QJQ, and LL were responsible for the final decision to submit the manuscript for publication.

## Acknowledgments

This work was supported by the National Basic Science Data Center ‘Interdisciplinary Brain Database for In vivo Population Imaging’ (ID-BRAIN to Xi-Nian Zuo).

## For the Lifespan Brain Chart Consortium (LBCC)

Chris Adamson^4,5^, Sophie Adler^6^, Aaron F. Alexander-Bloch^7,8,9^, Evdokia Anagnostou^10,11^, Kevin M. Anderson^12^, Ariosky Areces-Gonzalez^13,14^, Duncan E. Astle^15^, Bonnie Auyeung^16,17^, Muhammad Ayub^18,19^, Jong Bin Bae^20^, Gareth Ball^4,21^, Simon Baron-Cohen^17,22^, Richard Beare^4,5^, Saashi A. Bedford^17^, Vivek Benegal^23^, Richard A.I. Bethlehem^17,24^, Frauke Beyer^25^, John Blangero^26^, Manuel Blesa Cábez^27^, James P. Boardman^27^, Matthew Borzage^28^, Jorge F. Bosch-Bayard^29,30^, Niall Bourke^31^, Edward T. Bullmore^32^, Vince D. Calhoun^33^, Mallar M. Chakravarty^34,30^, Christina Chen^35^, Casey Chertavian^9^, Gaël Chetelat^36^, Yap S. Chong^37,38^, Aiden Corvin^39^, Manuela Costantino^40,41^, Eric Courchesne^42,43^, Fabrice Crivello^44^, Vanessa L. Cropley^45^, Jennifer Crosbie^46^, Nicolas Crossley^47,48^, Marion Delarue^36^, Richard Delorme^49,50^, Sylvane Desrivieres^51^, Gabriel Devenyi^52,53^, Maria A. Di Biase^45,54^, Ray Dolan^55,56,57^, Kirsten A. Donald^58,59^, Gary Donohoe^60^, Lena Dorfschmidt^7,8,9^, Katharine Dunlop^61^, Anthony D Edwards^62,63,64^, Jed T. Elison^65^, Cameron T. Ellis^12,66^, Jeremy A. Elman^67^, Lisa Eyler^68,69^, Damien A. Fair^65^, Paul C. Fletcher^70,71^, Peter Fonagy^72,73^, Carol E. Franz^74^, Lidice Galan-Garcia^75^, Ali Gholipour^76^, Jay Giedd^77,78^, John H. Gilmore^79^, David C. Glahn^80,81^, Ian M. Goodyer^32^, P. E. Grant^82^, Nynke A. Groenewold^83,59^, Shreya Gudapati^7,8,9^, Faith M. Gunning^84^, Raquel E. Gur^7,9^, Ruben C. Gur^7,85^, Christopher F. Hammill^46,86^, Oskar Hansson^87,88^, Trey Hedden^89,90^, Andreas Heinz^91^, Richard N. Henson^15,92^, Katja Heuer^93^, Jacqueline Hoare^94^, Bharath Holla^95,96^, Avram J. Holmes^97^, Hao Huang^98^, Jonathan Ipser^99^, Clifford R. Jack Jr^100^, Andrea P. Jackowski^101,102^, Tianye Jia^103,104,105^, David T. Jones^106,107^, Peter B. Jones^32,108^, Rene S Kahn^109,110^, Hasse Karlsson^111,112^, Linnea Karlsson^111,112^, Ryuta Kawashima^113^, Elizabeth A. Kelley^114^, Silke Kern^115,116^, Ki-Woong Kim^117,118,119,120^, Manfred G. Kitzbichler^32^, William S. Kremen^67^, François Lalonde^121^, Brigitte Landeau^36^, Jason Lerch^122,123,124^, John D. Lewis^125^, Jiao Li^126^, Wei Liao^126^, Conor Liston^127^, Michael V. Lombardo^128,17^, Jinglei Lv^129,45^, Travis T. Mallard^130^, Machteld Marcelis^131^, Samuel R. Mathias^80^, Bernard Mazoyer^44,132^, Philip McGuire^133^, Michael J. Meaney^134^, Andrea Mechelli^133^, Bratislav Misic^135^, Sarah E Morgan^136,137,138^, David Mothersill^139,140,141^, Cynthia Ortinau^142^, Rik Ossenkoppele^143,144^, Minhui Ouyang^98^, Lena Palaniyappan^145^, Leo Paly^36^, Pedro M. Pan^146,147^, Christos Pantelis^148,149,150^, Min Tae M. Park^151^, Tomas Paus^152,153^, Zdenka Pausova^46,154^, Deirel Paz-Linares^13,155^, Alexa Pichet Binette^156,157^, Karen Pierce^158^, Xing Qian^159^, Anqi Qiu^160^, Armin Raznahan^121^, Timothy Rittman^161^, Amanda Rodrigue^80^, Caitlin K. Rollins^162,163^, Rafael Romero-Garcia^32,164^, Lisa Ronan^32^, Monica D. Rosenberg^165^, David H. Rowitch^166^, Giovanni A. Salum^167,168^, Theodore D. Satterthwaite^169,7^, H. Lina Schaare^170,171^, Jenna Schabdach^7,8,9^, Russell J. Schachar^46^, Michael Schöll^172,173,174^, Aaron P. Schultz^175,176,81^, Jakob Seidlitz^7,8,9^, David Sharp^177,178^, Russell T. Shinohara^35,179^, Ingmar Skoog^115,116^, Christopher D. Smyser^180^, Reisa A. Sperling^175,181,81^, Dan J. Stein^182^, Aleks Stolicyn^183^, John Suckling^32,184^, Gemma Sullivan^27^, Benjamin Thyreau^113^, Roberto Toro^93,185^, Nicolas Traut^186,187^, Kamen A. Tsvetanov^161,188^, Nicholas B. Turk-Browne^12,189^, Jetro J. Tuulari^190,191,192^, Christophe Tzourio^193^, É tienne Vachon-Presseau^194,195,196^, Mitchell J. Valdes-Sosa^75^, Pedro A. Valdes-Sosa^197,198^, Sofie L. Valk^199^, Therese van Amelsvoort^200^, Simon N. Vandekar^201,202^, Lana Vasung^203^, Petra E VÉrtes^32,204^, Lindsay W. Victoria^84^, Sylvia Villeneuve^157,205,156^, Arno Villringer^25,206^, Jacob W. Vogel^169,7^, Konrad Wagstyl^207^, Yin-Shan Wang^1,208,209,210^, Simon K. Warfield^76^, Varun Warrier^211^, Eric Westman^212^, Margaret L. Westwater^32^, Heather C. Whalley^183,213^, Simon R. White^32,214^, A. Veronica Witte^206,25,215^, Ning Yang^1,208,209,210^, B.T. Thomas Yeo^216,217,218,219^, Hyuk Jin Yun^220^, Andrew Zalesky^221^, Heather J Zar^83,222^, Anna Zettergren^115^, Juan H. Zhou^159,223,216^, Hisham Ziauddeen^32,224,225^, Dabriel Zimmerman^7,8,9^, Andre Zugman^226,227,147^, Xi-Nian Zuo^1,208,209,228^

^4^Developmental Imaging, Murdoch Children’s Research Institute, Melbourne, Victoria, Australia

^5^Department of Medicine, Monash University, Melbourne, Victoria, Australia

^6^UCL Great Ormond Street Institute for Child Health, 30 Guilford St, Holborn, London WC1N 1EH

^7^Department of Psychiatry, University of Pennsylvania, Philadelphia, PA 19104

^8^Department of Child and Adolescent Psychiatry and Behavioral Science, The Children’s Hospital of Philadelphia, Philadelphia, PA 19104

^9^Lifespan Brain Institute, The Children’s Hospital of Philadelphia, Philadelphia, PA 19104

^10^Department of Pediatrics University of Toronto

^11^Holland Bloorview Kids Rehabilitation Hospital, Toronto, Canada

^12^Department of Psychology, Yale University, New Haven, CT, USA

^13^The Clinical Hospital of Chengdu Brain Science Institute, MOE Key Lab for NeuroInformation, University of Electronic Science and Technology of China, No. 2006, Xiyuan Ave., West Hi-Tech Zone, Chengdu, 611731, China

^14^University of Pinar del Río “Hermanos Saiz Montes de Oca”, Cuba

^15^MRC Cognition and Brain Sciences Unit, University of Cambridge, Cambridge UK

^16^Department of Psychology, School of Philosophy, Psychology and Language Sciences, University of Edinburgh, Edinburgh, United Kingdom

^17^Autism Research Centre, Department of Psychiatry, University of Cambridge, Cambridge, CB2 0SZ, UK

^18^Queen’s University, Department of Psychiatry, Centre for Neuroscience Studies, Kingston, Ontario, Canada

^19^University College London, Mental Health Neuroscience Research Department, Division of Psychiatry, London UK

^20^Department of Neuropsychiatry, Seoul National University Bundang Hospital, Seongnam, Korea

^21^Department of Paediatrics, University of Melbourne, Melbourne, Victoria, Australia

^22^Cambridge Lifetime Asperger Syndrome Service (CLASS), Cambridgeshire and Peterborough NHS Foundation Trust, Cambridge, United Kingdom

^23^Centre for Addiction Medicine, National Institute of Mental Health and Neurosciences (NIMHANS), Bengaluru, India 560029

^24^Brain Mapping Unit, Department of Psychiatry, University of Cambridge, Cambridge, CB2 0SZ, UK

^25^Department of Neurology, Max Planck Institute for Human Cognitive and Brain Sciences, Leipzig, 04103, Germany

^26^Department of Human Genetics, South Texas Diabetes and Obesity Institute, University of Texas Rio Grande Valley

^27^MRC Centre for Reproductive Health, University of Edinburgh, UK

^28^Fetal and Neonatal Institute, Division of Neonatology, Children’s Hospital Los Angeles, Department of Pediatrics, Keck School of Medicine, University of Southern California, Los Angeles, California USA

^29^McGill Centre for Integrative Neuroscience, Ludmer Centre for Neuroinformatics and Mental Health, Montreal Neurological Institute

^30^McGill University

^31^Department of Brain Sciences, Imperial College London, London UK & Care Research & Technology Centre, UK Dementia Research Institute

^32^Department of Psychiatry, University of Cambridge, Cambridge, CB2 0SZ, UK

^33^Tri-institutional Center for Translational Research in Neuroimaging and Data Science, Georgia State University, Georgia Institute of Technology, and Emory University, Atlanta, GA, USA

^34^Computational Brain Anatomy (CoBrA) Laboratory, Cerebral Imaging Centre, Douglas Mental Health University Institute

^35^Penn Statistics in Imaging and Visualization Center, Department of Biostatistics, Epidemiology, and Informatics, Perelman School of Medicine, University of Pennsylvania, Philadelphia, PA, USA

^36^Normandie Univ, UNICAEN, INSERM, U1237, PhIND “Physiopathology and Imaging of Neurological Disorders”, Institut Blood and Brain @ Caen-Normandie, Cyceron, 14000 Caen, France

^37^Singapore Institute for Clinical Sciences, Agency for Science, Technology and Research, Singapore

^38^Department of Obstetrics and Gynaecology, Yong Loo Lin School of Medicine, National University of Singapore, Singapore

^39^Department of Psychiatry, Trinity College, Dublin, Ireland

^40^Cerebral Imaging Centre, Douglas Mental Health University Institute, Verdun, Canada

^41^Undergraduate program in Neuroscience, McGill University, Montreal, Canada

^42^Department of Neuroscience, University of California, San Diego, San Diego, CA 92093, USA

^43^Autism Center of Excellence, University of California, San Diego, San Diego, CA 92037, USA

^44^Institute of Neurodegenerative Disorders, CNRS UMR5293, CEA, University of Bordeaux

^45^Melbourne Neuropsychiatry Centre, University of Melbourne, Melbourne, Australia

^46^The Hospital for Sick Children, Toronto, Canada

^47^Department of Psychiatry, School of Medicine, Pontificia Universidad Católica de Chile, Diagonal Paraguay 362, Santiago 8330077, Chile

^48^Department of Psychiatry, University of Oxford OX3 7JX

^49^Child and Adolescent Psychiatry Department, Robert DebrÉ University Hospital, AP-HP, F-75019, Paris France

^50^Human Genetics and Cognitive Functions, Institut Pasteur, F-75015, Paris France

^51^Social, Genetic and Developmental Psychiatry Centre, Institute of Psychiatry, Psychology & Neuroscience, King’s College London, London, United Kingdom

^52^Cerebral Imaging Centre, Douglas Mental Health University Institute, Montreal, QC, Canada, McGill Department of Psychiatry, Montreal, QC, Canada

^53^Department of Psychiatry, McGill University, Montreal, QC, Canada

^54^Department of Psychiatry, Brigham and Women’s Hospital, Harvard Medical School, Boston, Massachusetts, United States

^55^Max Planck UCL Centre for Computational Psychiatry and Ageing Research, University College London, London, UK

^56^Wellcome Centre for Human Neuroimaging, University College London, London, UK

^57^Wellcome Centre for Human Neuroimaging, 12 Queen Square, London WC1N 3AR

^58^Division of Developmental Paediatrics, Department of Paediatrics and Child Health, Red Cross War Memorial Children’s Hospital, Klipfontein Road/Private Bag, Rondebosch, 7700/7701, Cape Town, South Africa

^59^Neuroscience Institute, University of Cape Town, Cape Town, South Africa

^60^Center for Neuroimaging, Cognition & Genomics (NICOG), School of Psychology, National University of Ireland Galway, Galway, Ireland

^61^Weil Family Brain and Mind Research Institute, Department of Psychiatry, Weill Cornell Medicine

^62^Centre for the Developing Brain, King’s College London, London, UK

^63^Evelina London Children’s Hospital

^64^MRC Centre for Neurodevelopmental Disorders, London

^65^Institute of Child Development, Department of Pediatrics, Masonic Institute for the Developing Brain, University of Minnesota, Minneapolis, MN, United States

^66^Haskins Laboratories, New Haven, CT, USA

^67^Department of Psychiatry, Center for Behavior Genetics of Aging, University of California, San Diego, La Jolla, CA

^68^Desert-Pacific Mental Illness Research Education and Clinical Center, VA San Diego Healthcare, San Diego, CA, USA

^69^Department of Psychiatry, University of California San Diego, Los Angeles, CA, USA

^70^Department of Psychiatry, University of Cambridge, and Wellcome Trust MRC Institute of Metabolic Science, Cambridge Biomedical Campus, Cambridge, United Kingdom

^71^Cambridgeshire and Peterborough NHS Foundation Trust

^72^Department of Clinical, Educational and Health Psychology, University College London, London, UK

^73^Anna Freud National Centre for Children and Families, London UK

^74^Department of Psychiatry, Center for Behavior Genetics of Aging, University of California, San Diego, La Jolla, CA 92093

^75^Cuban Center for Neuroscience, La Habana, Cuba

^76^Computational Radiology Laboratory, Boston Children’s Hospital, Boston, MA 02115

^77^Department of Child and Adolescent Psychiatry, University of California, San Diego, San Diego, CA 92093, USA

^78^Department of Psychiatry, University of California San Diego, San Diego, CA, USA

^79^Department of Psychiatry, University of North Carolina, Chapel Hill, NC, USA

^80^Department of Psychiatry, Boston Children’s Hospital and Harvard Medical School, Boston, MA 02115

^81^Harvard Medical School, Boston, MA 02115

^82^Division of Newborn Medicine and Neuroradiology, Fetal Neonatal Neuroimaging and Developmental Science Center, Boston Children’s Hospital, Harvard Medical School, Boston, MA 02115, USA

^83^Department of Paediatrics and Child Health, Red Cross War Memorial Children’s Hospital, SA-MRC Unit on Child & Adolescent Health, University of Cape Town, South Africa

^84^Weill Cornell Institute of Geriatric Psychiatry, Department of Psychiatry, Weill Cornell Medicine

^85^Lifespan Brain Institute, The Children’s Hospital of Philadelphia, Philadelphia, PA 19105

^86^Mouse Imaging Centre, Toronto, Canada

^87^Clinical Memory Research Unit, Department of Clinical Sciences Malmö, Lund University, Malmö, Sweden

^88^Memory Clinic, Skåne University Hospital, Malmö, Sweden

^89^Department of Neurology, Icahn School of Medicine at Mount Sinai, New York, NY 10029, USA

^90^Athinoula A. Martinos Center for Biomedical Imaging, Department of Radiology, Massachusetts General Hospital, Harvard Medical School, Boston, MA 02129, USA

^91^Department of Psychiatry and Psychotherapy, Charite University Hospital Berlin, Berlin, Germany

^92^Department of Psychiatry, University of Cambridge, Cambridge, UK

^93^Institut Pasteur, UniversitÉ Paris CitÉ, UnitÉ de Neuroanatomie AppliquÉe et ThÉorique, F-75015 Paris, France

^94^Department of Psychiatry, University of Cape Town, Cape Town, South Africa

^95^Department of Integrative Medicine, NIMHANS, Bengaluru-560029, India

^96^Accelerator Program for Discovery in Brain disorders using Stem cells (ADBS), Department of Psychiatry, NIMHANS, Bengaluru-560029, India

^97^Department of Psychiatry, Brain Health Institute, Rutgers University, Piscataway, NJ, USA

^98^Department of Radiology, Children’s Hospital of Philadelphia and University of Pennsylvania, Philadelphia, PA 19104

^99^Department of Psychiatry and Mental Health, Clinical Neuroscience Institute, University of Cape Town

^100^Department of Radiology, Mayo Clinic, Rochester, MN 55905, USA

^101^Department of Psychiatry, Universidade Federal de São Paulo

^102^National Institute of Developmental Psychiatry, CNPq

^103^Institute of Science and Technology for Brain-Inspired Intelligence, Fudan University, Shanghai, 200433, China

^104^Key Laboratory of Computational Neuroscience and BrainInspired Intelligence (Fudan University), Ministry of Education, Shanghai, China

^105^Centre for Population Neuroscience and Precision Medicine (PONS), Institute of Psychiatry, Psychology and Neuroscience, SGDP Centre, King’s College London, London SE5 8AF, UK

^106^Department of Neurology, Mayo Clinic, Rochester, MN, USA

^107^Department of Radiology, Mayo Clinic, Rochester, MN, USA

^108^Cambridgeshire and Peterborough NHS Foundation Trust, Huntingdon, United Kingdom

^109^Department of Psychiatry, Icahn School of Medicine at Mount Sinai, New York, NY, USA

^110^Department of Psychiatry, Icahn School of Medicine, Mount Sinai, New York, USA

^111^Department of Clinical Medicine, Department of Psychiatry and Turku Brain and Mind Center, FinnBrain Birth Cohort Study, University of Turku and Turku University Hospital, Turku, Finland

^112^Centre for Population Health Research, Turku University Hospital and University of Turku, Turku, Finland

^113^Institute of Development, Aging and Cancer, Tohoku University, Seiryocho, Aobaku, Sendai 980-8575, Japan

^114^Queen’s University, Departments of Psychology and Psychiatry, Centre for Neuroscience Studies, Kingston, On- tario, Canada

^115^Neuropsychiatric Epidemiology Unit, Department of Psychiatry and Neurochemistry, Institute of Neuroscience and Physiology, the Sahlgrenska Academy, Centre for Ageing and Health (AGECAP) at the University of Gothen- burg, Sweden

^116^Region Västra Götaland, Sahlgrenska University Hospital, Psychiatry, Cognition and Old Age Psychiatry Clinic, Gothenburg, Sweden

^117^Department of Brain and Cognitive Sciences, Seoul National University College of Natural Sciences, Seoul, Re- public of Korea

^118^Department of Neuropsychiatry, Seoul National University Bundang Hospital, Seongnam, Republic of Korea

^119^Department of Psychiatry, Seoul National University College of Medicine, Seoul, Republic of Korea

^120^Department of Brain and Cognitive Science, Seoul National University College of Natural Sciences

^121^Section on Developmental Neurogenomics, Human Genetics Branch, National Institute of Mental Health, Bethesda, MD, USA

^122^Department of Medical Biophysics, University of Toronto, Toronto, ON, Canada

^123^Mouse Imaging Centre, The Hospital for Sick Children, Toronto, ON, Canada

^124^Wellcome Centre for Integrative Neuroimaging, FMRIB, Nuffield Department of Clinical Neuroscience, University of Oxford, Oxford, UK

^125^Montreal Neurological Institute, McGill University, Montreal, Canada

^126^The Clinical Hospital of Chengdu Brain Science Institute, University of Electronic Science and Technology of China, Chengdu 611731, China

^127^Department of Psychiatry and Brain and Mind Research Institute, Weill Cornell Medicine

^128^Laboratory for Autism and Neurodevelopmental Disorders, Center for Neuroscience and Cognitive Systems @UniTn, Istituto Italiano di Tecnologia, Rovereto, Italy

^129^School of Biomedical Engineering & Brain and Mind Centre, The University of Sydney, Sydney, NSW, Australia

^130^Department of Psychology, University of Texas, Austin, Texas 78712, USA

^131^Department of Psychiatry and Neuropsychology, School of Mental Health and Neuroscience, EURON, Maastricht University Medical Centre, PO Box 616, 6200 MD, Maastricht, the Netherlands; Institute for Mental Health Care Eindhoven (GGzE), Eindhoven, the Netherlands

^132^Bordeaux University Hospital

^133^Professor, Department of Psychosis Studies, Institute of Psychiatry, Psychology and Neuroscience, King’s College London, UK

^134^Ludmer Centre for Neuroinformatics and Mental Health, Douglas Mental Health University Institute, McGill University, Montreal, Quebec, Canada; Singapore Institute for Clinical Sciences, Singapore

^135^McConnell Brain Imaging Centre, Montreal Neurological Institute, McGill University, Montreal, QC H3A 2B4, Canada

^136^Department of Computer Science and Technology, University of Cambridge, Cambridge CB3 0FD, United King- dom

^137^Department of Psychiatry, University of Cambridge, Cambridge CB2 0SZ, United Kingdom

^138^The Alan Turing Institute, London, NW1 2DB

^139^Department of Psychology, School of Business, National College of Ireland, Dublin, Ireland

^140^School of Psychology & Center for Neuroimaging and Cognitive Genomics, National University of Ireland Galway, Galway, Ireland

^141^Department of Psychiatry, Trinity College Dublin, Dublin, Ireland

^142^Department of Pediatrics, Washington University in St. Louis, St. Louis, Missouri, United States

^143^Alzheimer Center Amsterdam, Department of Neurology, Amsterdam Neuroscience, Vrije Universiteit Amsterdam, Amsterdam UMC, Amsterdam, The Netherlands

^144^Lund University, Clinical Memory Research Unit, Lund, Sweden

^145^Robarts Research Institute & The Brain and Mind Institute, University of Western Ontario,London,Ontario,Canada

^146^Department of Psychiatry, Federal University of Sao Poalo (UNIFESP)

^147^National Institute of Developmental Psychiatry for Children and Adolescents (INPD), Brazil

^148^Melbourne Neuropsychiatry Centre, Department of Psychiatry, The University of Melbourne and Melbourne Health, Carlton South, Victoria, Australia

^149^Melbourne School of Engineering, The University of Melbourne, Parkville, Victoria, Australia

^150^Florey Institute of Neuroscience and Mental Health, Parkville, VIC, Australia

^151^Department of Psychiatry, Schulich School of Medicine and Dentistry, Western University, London, ON, Canada ^152^Department of Psychiatry, Faculty of Medicine and Centre Hospitalier Universitaire Sainte-Justine, University of Montreal, Montreal, Quebec, Canada

^153^Departments of Psychiatry and Psychology, University of Toronto, Toronto, ON, Canada

^154^Departments of Physiology and Nutritional Sciences, University of Toronto, Toronto, Canada

^155^Cuban Neuroscience Center, Havana, Cuba

^156^Department of Psychiatry, Faculty of Medicine, McGill University, Montreal, Qc, H3A 1Y2, Canada

^157^Douglas Mental Health University Institute, Montreal, Qc, H4H 1R3, Canada

^158^Department of Neurosciences, University of California, San Diego La Jolla, CA, USA

^159^Center for Sleep and Cognition, Yong Loo Lin School of Medicine, National University of Singapore, Singapore

^160^Department of Biomedical Engineering, The N.1 Institute for Health, National University of Singapore

^161^Department of Clinical Neurosciences, University of Cambridge, Cambridge UK

^162^Department of Neurology, Harvard Medical School

^163^Department of Neurology, Boston Children’s Hospital, Boston, MA 02115

^164^Instituto de Biomedicina de Sevilla (IBiS) HUVR/CSIC/Universidad de Sevilla, Dpto. de Fisiología MÉdica y Biofísica, Spain

^165^Department of Psychology, Neuroscience Institute, University of Chicago

^166^Department of Paediatrics and Wellcome-MRC Cambridge Stem Cell Institute, University of Cambridge, Hills Road, Cambridge, UK

^167^Department of Psychiatry, Universidade Federal do Rio Grande do Sul (UFRGS)

^168^National Institute of Developmental Psychiatry (INPD)

^169^Lifespan Informatics & Neuroimaging Center, University of Pennsylvania, Philadelphia, PA 19104

^170^Otto Hahn Group Cognitive Neurogenetics, Max Planck Institute for Human Cognitive and Brain Sciences, Leipzig, Germany

^171^Institute of Neuroscience and Medicine (INM-7: Brain and Behaviour), Research Centre Juelich, Juelich, Germany

^172^Wallenberg Centre for Molecular and Translational Medicine, University of Gothenburg, Gothenburg, Sweden

^173^Department of Psychiatry and Neurochemistry, University of Gothenburg, Sweden

^174^Dementia Research Centre, Queen’s Square Institute of Neurology, University College London, UK

^175^Harvard Aging Brain Study, Department of Neurology, Massachusetts General Hospital, Boston, MA 02114

^176^Athinoula A. Martinos Center for Biomedical Imaging, Department of Radiology, Massachusetts General Hospital, Charlestown, MA 02129, USA

^177^Department of Brain Sciences, Imperial College London, London UK

^178^Care Research & Technology Centre, UK Dementia Research Institute

^179^Center for Biomedical Image Computing and Analytics, Department of Radiology, Perelman School of Medicine, University of Pennsylvania, Philadelphia, PA, USA

^180^Departments of Neurology, Pediatrics, and Radiology, Washington University School of Medicine, St. Louis, United States

^181^Center for Alzheimer Research and Treatment, Department of Neurology, Brigham and Women’s Hospital, Boston, MA 02115

^182^SA MRC Unit on Risk & Resilience in Mental Disorders, Dept of Psychiatry and Neuroscience Institute, University of Cape Town, Cape Town, South Africa

^183^Division of Psychiatry, Centre for Clinical Brain Sciences, University of Edinburgh, UK

^184^Cambridge and Peterborough Foundation NHS Trust

^185^UniversitÉ de Paris, Paris, France

^186^Department of Neuroscience, Institut Pasteur, Paris, France

^187^Center for Research and Interdisciplinarity (CRI), UniversitÉ Paris Descartes, Paris, France

^188^Department of Psychology, University of Cambridge, Cambridge, UK

^189^Wu Tsai Institute, Yale University, New Haven, CT, USA

^190^Department of Clinical Medicine, Department of Psychiatry, FinnBrain Birth Cohort Study, University of Turku, Turku, Finland

^191^Department of Clinical Medicine, University of Turku, Turku Finland

^192^Turku Collegium for Science, Medicine and Technology, University of Turku, Turku, Finland

^193^Univ. Bordeaux, Inserm, Bordeaux Population Health Research Center, U1219, CHU Bordeaux, F-33000 Bor- deaux, France

^194^Faculty of Dental Medicine and Oral Health Sciences, McGill University, Montreal, Qc, H3A 1G1, Canada

^195^Faculty of Dentistry, McGill University, Montreal, Qc, H3A 1G1, Canada

^196^Alan Edwards Centre for Research on Pain (AECRP), McGill University, Montreal, Qc, H3A 1G1, Canada

^197^Joint China-Cuba Lab,University of Electronic Science and Technology, Chengdu China/Cuban Center for Neuroscience, La Habana, Cuba

^198^University of Electronic Science and Technology of China/Cuban Center for Neuroscience

^199^Institute for Neuroscience and Medicine 7, Forschungszentrum Juelich; Max Planck Institute for Human Cognitive and Brain Sciences

^200^Department of Psychiatry & Neurosychology, Maastricht University, Maastricht, The Netherlands

^201^Department of Biostatistics, Vanderbilt University, Nashville, Tennessee, USA

^202^Department of Biostatistics, Vanderbilt University Medical Center, Nashville, Tennessee, USA

^203^Division of Newborn Medicine, Fetal Neonatal Neuroimaging and Developmental Science Center, Department of Pediatrics, Boston Children’s Hospital, Boston, MA 02115

^204^The Alan Turing Institute, London NW1 2DB, UK

^205^McConnell Brain Imaging Center, Montreal Neurological Institute, McGill University, Montreal, Quebec, Canada

^206^Clinic for Cognitive Neurology, University of Leipzig Medical Center, Leipzig, 04103, Germany

^207^Wellcome Centre for Human Neuroimaging, Institute of Neurology, University College London, WC1N 3AR

^208^Developmental Population Neuroscience Research Center, IDG/McGovern Institute for Brain Research, Beijing Normal University, Beijing 100875, China

^209^National Basic Science Data Center, Beijing 100190, China

^210^Research Center for Lifespan Development of Brain and Mind, Institute of Psychology, Chinese Academy of Sciences, Beijing 100101, China

^211^Department of Psychiatry, University of Cambridge, Cambridge, CB2 0SZ, UK

^212^Division of Clinical Geriatrics, Center for Alzheimer Research, Department of Neurobiology, Care Sciences and Society, Karolinska Institutet, Stockholm, Sweden

^213^Generation Scotland, University of Edinburgh

^214^MRC Biostatistics Unit, University of Cambridge, Cambridge, England

^215^Faculty of Medicine, CRC 1052 ‘Obesity Mechanisms’, University of Leipzig, Leipzig, 04103, Germany

^216^Department of Electrical and Computer Engineering, National University of Singapore, Singapore

^217^Centre for Sleep & Cognition and Centre for Translational MR Research, Yong Loo Lin School of Medicine, National University of Singapore, Singapore

^218^N.1 Institute for Health & Institute for Digital Medicine, National University of Singapore, Singapore

^219^Integrative Sciences and Engineering Programme (ISEP), National University of Singapore, Singapore

^220^Fetal Neonatal Neuroimaging and Developmental Science Center, Division of Newborn Medicine, Boston Children’s Hospital, Harvard Medical School, Boston, MA 02115, USA

^221^Melbourne Neuropsychiatry Centre, University of Melbourne, Melbourne, Australia; Department of Biomedical Engineering, University of Melbourne, Melbourne, Australia

^222^SAMRC Unit on Child & Adolescent Health, University of Cape Town, South Africa

^223^Center for Translational Magnetic Resonance Research, Yong Loo Lin School of Medicine, National University of Singapore, Singapore

^224^Wellcome Trust-MRC Institute of Metabolic Science, University of Cambridge, Cambridge, CB2 0SZ

^225^Cambridgeshire and Peterborough Foundation Trust, Cambridge, CB21 5EF

^226^National Institute of Mental Health (NIMH), National Institutes of Health (NIH), Bethesda, Maryland, USA

^227^Department of Psychiatry, Escola Paulista de Medicina, São Paulo, Brazil

^228^Research Center for Lifespan Development of Brain and Mind, Institute of Psychology, Chinese Academy of Sciences, Beijing 100101, China ; Key Laboratory of Brain and Education, School of Education Science, Nanning Normal University, Nanning 530001, China

## For the Chinese Color Nest Consortium (CCNC)

Xi-Nian Zuo^1,2,3,4,7,18^, Ning Yang^1,2,3,4^, Zhe Zhang^5^, Ye He^6^, Hao-Ming Dong^1,4,8^, Lei Zhang^9^, Xing-Ting Zhu^2^, Xiao-Hui Hou^7^, Yin-Shan Wang^1,3^, Quan Zhou^1,2,3^, Zhu-Qing Gong^1,2,3^, Li-Zhi Cao^2,4^, Ping Wang^1^, Yi- Wen Zhang^2^, Dan-Yang Sui^20^, Ting Xu^21^, Gao-Xia Wei^2,4^, Zhi Yang^2,4^, Lili Jiang^2,4^, Hui-Jie Li^2,4^, Ting-Yong Feng^11^, Antao Chen^11^, Jiang Qiu^11^, Xu Chen^11^, Xun Liu^2,4^, Ke Zhao^2,4^, Yi Du^2,4^, Min Bao^2,4^, Yuan Zhou^2,4^, Yan Zhuo^14^, Zhentao Zuo^14^, Li Ke^1^, Fei Wang^22^, Zhi-Xiong Yan^7^, Xue-Quan Su^7^, F.Xavier Castellanos^23^, Michael Peter Milham^21,24^, Yu-Feng Zang^25^

^1^State Key Laboratory of Cognitive Neuroscience and Learning, Beijing Normal University, Beijing, 100875, China.

^2^Department of Psychology, University of Chinese Academy of Sciences, Beijing, 100049, China.

^3^Developmental Population Neuroscience Research Center, International Data Group/McGovern Institute for Brain Research, Beijing Normal University, Beijing, 100875, China.

^4^Key Laboratory of Behavioural Science, Institute of Psychology, Chinese Academy of Sciences, Beijing, 100101, China.

^5^College of Education, Hebei Normal University, Shijiazhuang, 050024, China.

^6^School of Artificial Intelligence, Beijing University of Posts and Telecommunications, Beijing, 100876, China.

^7^Laboratory of Cognitive Neuroscience and Education, School of Education Science, Nanning Normal University, Nanning, 530299, China.

^8^Changping Laboratory, Beijing, 102206, China.

^9^School of Government, Shanghai University of Political Science and Law, Shanghai, 201701, China.

^10^School of Psychology, Research Center for Exercise and Brain Science, Shanghai University of Sport, Shanghai, 200438, China.

^11^Faculty of Psychology, Southwest University, Chongqing, 400715, China.

^12^NYU-ECNU Institute of Brain and Cognitive Science at New York University Shanghai, Shanghai, 200062, China.

^13^Faculty of Arts and Science, New York University Shanghai, Shanghai, 200122, China.

^14^State Key Laboratory of Brain and Cognitive Science, Institute of Psychology, Chinese Academy of Sciences, Bei- jing, 100101, China.

^15^School of Psychology and Cognitive Science, East China Normal University, Shanghai, 200062, China.

^16^Department of Psychology, Renmin University of China, Beijing, 100872, China.

^17^Beijing Key Laboratory of Learning and Cognition, School of Psychology, Capital Normal University, Beijing, 100048, China.

^18^School of Education, Hunan University of Science and Technology, Hunan Xiangtan, 411201, China.

^19^National Basic Science Data Center, Beijing, 100190, China.

